# Effects of socio-economic factors on elementary school student COVID-19 infections in Ontario, Canada

**DOI:** 10.1101/2022.02.04.22270413

**Authors:** Prachi Srivastava, Tsz Tan Lau, Daniel Ansari, Nisha Thampi

## Abstract

**Background:** The prevalence of SARS-CoV-2 infections in Ontario is disproportionately concentrated in areas with lower-income and racialized groups. We examined whether school-level and area-level socio-economic factors were associated with elementary school student infections in Ontario.

**Methods:** We performed multi-level modeling analyses using data from the Ministry of Education on school-based infections in Ontario in the 2020-21 school year and on school-level demographics, the Ontario Marginalization Index, and census data to estimate the variability of the cumulative incidence of SARS-CoV-2 infections amongst elementary school students attributable to individual schools (school level, Level 1) and forward sortation areas (FSAs) of schools (area level, Level 2). We explored whether socio-economic factors within individual schools and/or factors common to schools within FSAs predicted the incidence of elementary school student infections.

**Results:** At the school level, the proportion of students from low-income households within a school was positively related with the cumulative incidence of SARS-CoV-2 elementary school student infections (*β* = .083, *p* < 0.001). At the area level, the dimensions of FSA marginalization were significantly related with cumulative incidence. Ethnic concentration (*β* = .454, *p* < 0.001), residential instability (*β* = .356, *p* < 0.001), and material deprivation (*β* = .212, *p* < 0.001) were positively related. Area-related variables were more likely to explain variance in cumulative incidence than school-related variables (58% versus 1%, respectively).

**Interpretation:** Socio-economic characteristics of the geographic location of schools were more important in determining the cumulative incidence of SARS-CoV-2 elementary school student infections than individual school characteristics. Given inequitable effects of protracted education disruption, schools in marginalized areas should be prioritized for infection prevention measures and education continuity and recovery plans.

## INTRODUCTION

The prevalence of SARS-CoV-2 infections has been disproportionately concentrated in neighbourhoods with lower-income and racialized groups in Canada,^1^ and in its largest city, Toronto, Ontario.^2^ School closures elsewhere have been disproportionately experienced in marginalized areas.^3^ As schools are nested within specific geographic areas, this raises critical health and social equity concerns affecting children and youth. However, analyses of the potential differential effects of socio-economic factors on school infections in Canada are lacking. Specifically, it is unclear whether school-related infections were more likely to occur in schools and/or areas with aggravated indicators of disadvantage.

The education sector is in need of urgent equity-focused analysis in view of the scale of pandemic-related disruption and known negative effects.^4,5,6^ Given the well-established literature on the nested nature of schools in specified geographic areas and resulting equity effects,^7,8,9^ we sought to understand the relationship between school-level and area-level factors on the cumulative incidence of laboratory-confirmed SARS-CoV-2 student infections within and across publicly funded elementary schools in Ontario for the 2020-21 school year.

Ontario has the largest cohort of elementary and secondary students in Canada. Publicly funded schools claim 94% of all enrolment in the province.^10,11^ Furthermore, Ontario had the longest period of school closures in the country, at an average of 26 weeks between March 2020 and June 2021.^12^

The COVID-19 School Dashboard was developed as a data visualization platform to enable analysis between systems-level education data and school-related infections.^13^ The underlying data integrate all officially reported cases in publicly funded schools in Ontario, with school demographic and geo-location data. Knowing the geographic distribution of all school cases, we investigated the effects of school- and area-level socio-economic factors on the cumulative incidence of laboratory-confirmed SARS-CoV-2 elementary school student infections.

## METHODS

### School Selection

We focused on elementary schools to minimize contextual variation in pedagogical strategies and associated face-to-face interactions. During periods that Ontario schools were open for in-person instruction in 2020-21, elementary schools typically instituted full-day face-to-face classes during the entire week. This was in contrast to an adapted instructional model in secondary schools that employed a combination of reduced and alternative days of in-person and virtual instruction. We could not include private schools because neither data on private school-related infections nor on private school demographics were publicly released.

We defined the geographic area as the forward sortation area (FSA), i.e., the first three characters of the postal code. In principle, catchment areas structure how students are sorted into public schools in Ontario. Ideally, we would have had access to school catchment boundaries, however, they were not publicly available for all schools and school boards. The FSA was the most comparable unit available. The final sample consisted of 3994 publicly funded elementary schools across 491 FSAs in Ontario.

Specifically, we were interested in ascertaining whether the proportion of students from marginalized socio-economic backgrounds in any individual school had an effect on the cumulative incidence of student infections in that school (Level 1). We further sought to determine whether the proportion of households from marginalized backgrounds in a defined geographic area had an effect on the cumulative incidence of elementary school student infections across schools in that area (Level 2). Thus, we can discern whether socio-economic factors of the student body in a school had an effect on the cumulative incidence of student infections within that school, and also, whether schools in areas with relatively marginalized populations had higher cumulative incidences of student infections as compared to schools in other areas.

### Data Sources and Variables

We used the latest relevant publicly available data at the time of analysis. We obtained data on laboratory-confirmed cases of SARS-CoV-2 school-related infections and school administrative and student demographic data via the Government of Ontario Open Data Directive.^14^ We used the *Schools with Recent COVID-19 Cases (April 27, 2021 update)* dataset to extract laboratory-confirmed cases of SARS-CoV-2 in schools.^15^ The latest school-level demographic data were available for the 2019-20 school year, which we extracted from the *School Information and Student Demographics (June 29, 2021 update)* dataset.^16^ We used the Ontario Marginalization Index (ON-Marg) data (based on the 2016 census) for indicators of socio-economic marginalization at the area level.^17^ Finally, we obtained data on FSA population size from the 2016 census conducted by Statistics Canada.^18^

The dependent variable is the cumulative incidence of laboratory-confirmed elementary school student SARS-CoV-2 infections in the 2020-21 school year.^19,20^ The choice of school-level predictors was guided by the literature on pandemic school closures and education inequities,^21,22^ and limited to what was available in the school demographic dataset. School-level predictors (Level 1) were: proportion of low-income households, language mismatch (mismatch of student first language with school medium of instruction), and low parental education.^23^ The percentage of the student body that was recent immigrant was available but highly correlated with language mismatch, thus excluded. The school demographic dataset did not have race-based data.

Area-level predictors (Level 2) were the population of the FSA, the FSA average of the three school-level predictors above, and the four marginalization indicators from ON-Marg (i.e., residential instability, material deprivation, dependency, and ethnic concentration). Area means of the three school-level predictors were also considered as potential predictors. However, the means were highly correlated with ON-Marg variables raising multicollinearity concerns, and thus, excluded from the final model (Appendix A).

### Dependent Variable

#### Cumulative Incidence of Student SARS-CoV-2 Infections

The number of confirmed SARS-CoV-2 elementary and secondary school infections was reported every weekday by the Ontario Ministry of Education. Case counts were disaggregated in the original dataset as student, staff, and unidentified. This analysis includes elementary student cases only.

We inferred and computed the cumulative number of student cases, owing to the structure of the active cases dataset. The dataset included the total number of active cases per day but did not specify data on new cases. We computed new cases as additional cases of student infections on a given day when compared to the previous day’s total. We computed the cumulative number of new cases over the complete span of recorded time in the dataset (11 September 2020 to 15 April 2021).^24^ The computed cumulative case count was divided by the number of enrolled students per school and multiplied by 1000.^25,26^ The result reflects the number of cases per 1000 students in the school.

### School-level (Level 1) Predictors

#### Low-income Households

We extracted the proportion of students in a school belonging to low-income households from the school demographic dataset. This was defined in the dataset as the percentage of students in the school from households with income below the after-tax low-income measure threshold (LIM-AT) as defined by Statistics Canada. Each school estimated this proportion by using postal code data of the student body and cross-referencing it with income data from the 2016 census.^27,28^

#### Language Mismatch

We calculated language mismatch as the percentage of the student body whose first language did not match the medium of instruction at the school (i.e., percentage of students whose first language was not English in English-medium schools; percentage of students whose first language was not French in French-medium schools). Data on first language of the student body and the medium of instruction (English or French) were extracted from the school demographic dataset.

#### Low Parental Education

The proportion of students in a school from households with low parental education was extracted from the school demographic dataset. It was defined in the dataset as the percentage of students with parents who did not have a degree, diploma, or certificate.

### Area-level (Level 2) Predictors

#### FSA Population Size

We included all 491 of the 513 FSAs in Ontario containing publicly funded elementary schools.^29^ We extracted the population size for the relevant FSAs from the 2016 census.^30^

#### Ontario Marginalization Index

The ON-Marg is an area-based index that combines a number of demographic indicators into four dimensions of marginalization: residential instability, material deprivation, dependency, and ethnic concentration (Appendix B, description of constituent items).

### Analytic Methods

Analyses were performed using multi-level models with schools (school level, Level 1) nested in FSAs (area level, Level 2). We estimated two multi-level models (Appendix C, model specifications; Appendix D, additional analyses). The multi-level data structure comprised all 3994 elementary schools nested in 491 FSAs. All predictors were standardized (M = 0, S.D. = 1) prior to analysis to remove non-essential multi-collinearity.^31^ All analyses were conducted using Mplus 8.3,^32^ with the maximum likelihood estimator with robust standard errors.^33^ We report sensitivity tests of how the outcome variable was operationalized (cumulative incidence vs. cumulative cases) in Appendix E.

Model 1 was an intercept-only model estimating the variability of the cumulative incidence of SARS-CoV-2 elementary school student infections at the school level (Level 1) and the area level (Level 2). This enables us to calculate the intra-class correlation (ICC), i.e., the percentage of total variability in the cumulative incidence of student infections that can be accounted for by the geographic area (i.e., FSA) in which schools are located. Consequently, the higher the ICC, the more likely that two schools within the same geographic area will have similar numbers of cumulative incidences of student infections. This information is pertinent as it informs policymakers the degree to which elementary school student infections may be linked with the geographic location of the school.

Model 1 explores the degree to which variability of the cumulative incidence of SARS-CoV-2 elementary school student infections may be attributable to the individual schools or to geographic areas. Model 2 explores the degree to which school and area characteristics may be predictive of this variability. To this end, Model 2 included predictors at the school and area levels. At the school level, low-income households, language mismatch, and low parental education were used as predictors. FSA population, residential instability, material deprivation, dependency, and ethnic concentration were used as area-level predictors.

## RESULTS

### Descriptive Statistics

Table 1 presents characteristics of the Ontario public elementary school student population. All independent variables deviate moderately from normality (skewness < 2, kurtosis < 7).^34^ However, we observe high kurtosis for the cumulative incidence of student infections. This indicates that the cumulative incidence of student infections in most schools centered around the mean (5.36), with large fluctuations at extreme values.^35^

**Table 1.** Characteristics of the Ontario public elementary school student population.

|  | Mean | Median | SD | Skew | Kurtosis |
| --- | --- | --- | --- | --- | --- |
| **Overall Sample** |  |  |  |  |  |
| Cumulative incidence SARS-CoV-2 elementary school student infections (per 1000 students) | 5.36 | 2.70 | 8.10 | 3.52 | 24.45 |
| Low-income households (%) | 18.89 | 15.00 | 11.16 | 1.15 | 1.39 |
| Low parental education (%) | 6.03 | 5.00 | 6.72 | 2.11 | 8.18 |
| Language mismatch (%) | 23.10 | 15.00 | 23.11 | 1.21 | -0.16 |
| Population size of FSA | 38446.5 | 33027.00 | 23188.14 | 1.12 | 1.16 |
| Residential instability* | -0.22 | -0.32 | 0.57 | 0.85 | 1.28 |
| Material deprivation* | -0.18 | -0.24 | 0.5 | 0.67 | 0.71 |
| Dependency* | -0.06 | -0.07 | 0.42 | 0.24 | 0.82 |
| Ethnic concentration* | 0.3 | -0.04 | 1.01 | 1.14 | 0.25 |

\*: The four ON-Marg variables reflect factor scores constructed from principal component analysis.<sup>36</sup> Factor scores are standardized (Mean = 0, SD = 1) when the full Canada index is used. Observed deviations are due to aggregating ON-Marg score values to the FSA unit and using only factor scores from Ontario.

### Multi-level Models

Table 2 presents estimated Models 1 and 2. Model 1 tests whether there may be meaningful differences in the cumulative incidence of SARS-CoV-2 elementary school student infections across FSAs. Results indicate there were indeed meaningful between-FSA differences in the cumulative incidence. Variability at the area level (ICC) accounts for 15.5% of total variability. This suggests that there is modest between-area variability of school student infections, and that approximately 16% school variability of student infections can be explained by the geographic location of the school.

**Table 2.** Multi-level Models.

|  | **Model 1** | **Model 2** |
| --- | --- | --- |
|  | **Beta (S.E.)** | **Beta (S.E.)** |
| Intercept | 1.688(.111)*** | 1.921(.153)*** |
| **School-level (Level-1) Predictors** |  |  |
| Low-Income Households | - | .083(.024)*** |
| Language Mismatch | - | .035(.020) |
| Low Parental Education | - | .029(.021) |
| **Area-level (Level-2) Predictors** |  |  |
| Population of FSA | - | -.043(.047) |
| Residential Instability | - | .356(.060)*** |
| Material Deprivation | - | .212(.066)*** |
| Dependency | - | -.204(.056)*** |
| Ethnic Concentration | - | .454(.061)*** |
| **Random Effects** |  |  |
| Variance within Schools | 55.690(5.557)*** | 54.801(5.463)*** |
| Variance between FSAs | 10.242(1.403)*** | 4.715(0.750)*** |
| ICC<sub>school</sub> | .845 | .921 |
| ICC<sub>FSA</sub> | .155 | .079 |
| R<sup>2</sup><sub>school</sub> | - | .012 |
| R<sup>2</sup><sub>FSA</sub> | - | .576 |

\* $p < 0.5$ , \*\* $p < 0.01$ , \*\*\* $p < 0.001$ . Standardized Coefficients are presented.

Model 2 examines whether socio-economic factors and dimensions of marginalization may predict the cumulative incidence of SARS-CoV-2 elementary school student infections. Results show that at the school level (Level 1), the proportion of low-income households is significant and positively related with cumulative incidence (*β* = .083, *p* < 0.001). This suggests that when between-FSA variability is controlled for, a higher proportion of low-income households is related with higher cumulative incidence of student infections within the school. At the school level (Level 1), we found the proportion of low-income households within the school to predict the cumulative incidence of SARS-CoV-2 infections among elementary school students. School-level variables accounted for approximately 1% of school-level variability.

At the area level (Level 2), all four dimensions of marginalization were statistically significant. Whilst ethnic concentration (*β* = .454, *p* < 0.001), residential instability (*β* = .356, *p* < 0.001), and material deprivation (*β* = .212, *p* < 0.001) were positively related with the cumulative incidence of student infections, dependency (*β* = −.204, *p* < 0.001) was negatively related. This suggests that, on average, schools in FSAs with relatively higher levels of ethnic concentration, residential instability, and material deprivation would have higher cumulative incidence of student infections, while schools in FSAs with higher dependency would have lower cumulative incidences. The association with marginalization indicators was strong, accounting for 58% of variability at the area level.

## INTERPRETATION

Our analyses uncovered several important findings. First, a substantial proportion of variation in the cumulative incidence of SARS-CoV-2 elementary school student infections can be explained by the geographic location of schools. Second, the proportion of low-income households in individual schools was associated with the cumulative incidence of elementary school student infections, albeit weakly. Third, marginalization was strongly associated with geographic variability in the cumulative incidence of elementary school student infections.

Due to catchment restrictions, public elementary schools tend to draw students from local areas. Thus, variability in school infections could likely be attributable to the characteristics of individual school populations, as well as to the area in which schools are situated. As such, school infections may be predicted by variables unique to individual schools (e.g., proportion of students from low-income households or other socio-economic factors), and/or factors characterizing the area where schools are located (e.g., proportion of households from marginalized backgrounds within that area). From a policy perspective, understanding which level of analysis is most predictive can allow for greater precision in implementing school-based infection prevention measures, and education continuity and recovery strategies.

A multi-levelling modelling approach afforded us the opportunity to statistically determine which of the multiple school- and area-level factors predicted infections, by explicitly considering the nested nature of schools within particular geographic areas. The study strongly suggests that area characteristics, rather than factors unique to individual schools may have driven the cumulative incidence of SARS-CoV-2 elementary school student infections. Our results show that most of the variance in infections can be explained by area-level predictors.

Schools in areas with households experiencing more housing instability and material deprivation, and with higher concentrations of ethnic minorities were associated with higher cumulative incidence of elementary school student infections. Our findings are consistent with public health reporting in Ontario. Among school-aged SARS-CoV-2 cases, the most diverse neighbourhoods had rates that were approximately 3.5 times higher than those in the least diverse neighbourhoods, and rates were 1.6 times higher in the most deprived compared to the least deprived neighbourhoods.^37^ Results of our analysis show the effects of the embeddedness of schools within communities. Thus, student susceptibility to SARS-CoV-2 must be viewed through a social determinant framework for public health interventions to prevent onward transmission in schools and their communities.^38^

Finally, increasing inequities in prevalence have also been noted over time, and associated with households with lower incomes, crowding, and essential workers.^39,40^ Household size has been shown to be associated with increased odds of a positive test result for SARS-CoV-2, likely related to more intense exposure compared to other settings.^41^ Moreover, households had increased risk of infection associated with increasing number of children in the household.^42^ Interestingly, in our analysis, schools in areas with higher dependency (i.e., defined as households with individuals aged 65 and older, 14 and younger, and/or adults not participating in the labour force) had relatively lower cumulative incidences of student infections. This warrants further study to determine potential protective factors associated with dependency, e.g., reduced mobility and contacts outside the home, higher instances of virtual learning, and vaccine eligibility.

### Implications and Future Directions

There are equity-related implications of the analysis. While school closure data were not available, the implications of higher cumulative incidence of student infections in schools in marginalized areas is that those schools and school populations would have likely sustained relatively more disruption due to isolation measures at the individual level (i.e., students excluded), classroom level (i.e., classes isolated), and school level (i.e., school closures), than schools in other areas.^43^ This raises concerns. Systematic reviews of studies on COVID-19-related school closures indicate significant learning loss even with the provision of emergency virtual instruction, the bulk of studies showing greater losses among students from lower-income and other marginalized groups.^44,45^ In addition to learning loss, an earlier systematic review of extended education disruption during coronavirus outbreaks shows substantial well-documented harms, such as aggravated mental health effects and protection and social welfare concerns, disproportionately borne by socially disadvantaged groups.^46^ Thus, from a policy perspective, our findings suggest that schools in more marginalized areas should be prioritized for resources to reduce the risk of infection and associated education disruption.

### Limitations

Publicly available datasets do not include data on localized school closures instituted by public health units or by school boards, or on the numbers of days individual schools were closed or classes were in isolation. From a methodological perspective, this inhibits a more fine-grained computation of the cumulative incidence variable. The lack of publicly available race-based school-level data underscores the need for these data to be included in the school demographic dataset. Nonetheless, the datasets we used were the most complete we could publicly obtain and are consistent with data used by policymakers for decision-making. Similarly, we lacked catchment boundaries, thus we used marginalization data for FSAs. This is commonly employed for public health analyses. However, as catchment boundaries are stricter than large FSAs, it may be that our model is more conservative on the extent of variability predicted by area-level socio-economic factors. The exclusion of private schools in official data also likely leads to an underestimation.

## CONCLUSIONS

The socio-economic characteristics of the geographic location of schools were more strongly associated with the cumulative incidence of elementary school student infections than individual school characteristics. Elementary schools in marginalized areas in Ontario were more negatively affected. Schools with a higher proportion of students from lower-income households had a higher cumulative incidence of SARS-CoV-2 student infections in the 2020-21 school year, although this relationship was relatively weak. In lay terms, it mattered more, and very strongly, where schools were located than the socio-economic characteristics of individual school populations. Given the inequitable effects of protracted school closures, schools in more marginalized areas should be prioritized for infection prevention and mitigation measures, and in education continuity and recovery plans.

## Data Availability

All data used are available online at https://dataverse.scholarsportal.info/dataverse/COVID19SchoolInfections

## ACKNOWLEDGMENTS

The authors would like to thank Stefan Baral, Kate H. Choi, Amy Greer, and Peter J. Taylor for their inputs and comments.

## Appendix A. Multi-collinearity

Examining the correlations of area level (Level 2) predictors suggests that Level 2 aggregates of school characteristics were highly correlated with the four dimensions of ON-Marg. These high correlations raised concerns of multi-collinearity. Variance inflation factors were calculated (Table A1).

The FSA average of low-income households was correlated with material deprivation at 0.792. The FSA average of language mismatch was correlated with ethnic concentration at 0.878. The variance inflation factors were high for low-income households, language mismatch, material deprivation, and ethnic concentration (∼5). Furthermore, it is unlikely that schools exert a normative effect on schools in the same FSA (e.g., schools affecting how other schools in the FSA operated). As such, Level-2 aggregates of school characteristics may be substitutions of FSA characteristics. Given these considerations, the Level 2 aggregates of school characteristics were not included in Model 2.

**Table A1.** Correlation Matrix of Level-2 Variables.

|  | 1. | 2. | 3. | 4. | 5. | 6. | 7. | 8. |
| --- | --- | --- | --- | --- | --- | --- | --- | --- |
| 1. Low-Income Households | 1.00 |  |  |  |  |  |  |  |
| 2. Low Parental Education | 0.60 | 1.00 |  |  |  |  |  |  |
| 3. Language Mismatch | 0.39 | 0.11 | 1.00 |  |  |  |  |  |
| 4. Residential Instability | 0.61 | 0.23 | 0.12 | 1.00 |  |  |  |  |
| 5. Material Deprivation | 0.79 | 0.71 | 0.15 | 0.36 | 1.00 |  |  |  |
| 6. Dependency | 0.20 | 0.17 | -0.40 | 0.17 | 0.31 | 1.00 |  |  |
| 7. Ethnic Concentration | 0.38 | 0.12 | 0.88 | 0.06 | 0.21 | -0.52 | 1.00 |  |
| 8. Population of FSA | -0.16 | -0.08 | 0.18 | -0.31 | -0.16 | -0.28 | 0.28 | 1.00 |
| Variance Inflation Factor | 5.62 | 2.14 | 4.96 | 1.99 | 4.25 | 2.01 | 6.52 | 1.26 |

## Appendix B. Item Description of ON-Marg Variables

**Table B1.** On-Marg Variable Descriptions.

| Dimension | Definition | Indicators |
| --- | --- | --- |
| Residential Instability | Area concentration of people who experience family or housing instability | Proportion of the population living alone<br>Proportion of the population who are not youth (age 5-15)<br>Average number of persons per dwelling<br>Proportion of dwellings that are apartment buildings<br>Proportion of the population who are single/divorced/widowed<br>Proportion of dwellings that are not owned<br>Proportion of the population who moved during the past 5 years |
| Material Deprivation | Area concentration of individual and communities with inability to access and attain basic material needs | Proportion of the population aged 20+ without a high-school diploma<br>Proportion of families who are lone parent families<br>Proportion of total income from government transfer payments for population aged 15+<br>Proportion of the population aged 15+ who are unemployed<br>Proportion of the population considered low-income<br>Proportion of households living in dwellings that are in need of major repair |
| Dependency | Area concentration of individuals who do not have income from employment | Proportion of the population who are aged 65 and older<br>Dependency ratio (total population 0-14 and 65+ / total population 15 to 64)<br>Proportion of the population not participating in labour force (aged 15+) |
| Ethnic Concentration | Area concentration of people who belong to a minority group or have recently moved to Canada | Proportion of the population who are recent immigrants (arrived in the past 5 years)<br>Proportion of the population who self-identify as a visible minority |

## Appendix C. Model Specification

Details on model specifications are found below.

Model 1 was an intercept-only model with no predictors but allowed for the estimation of variability in the cumulative incidence of SARS-CoV-2 student infections at Level 1 (school) and Level 2 (FSA). Following Raudenbush & Bryk (Raudenbush & Bryk, 2002), Model 1 is as follows:

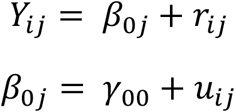

Where *Y*_*ij*_ refers to cumulative incidence of SARS-CoV-2 student infections for the i^th^ publicly funded elementary school in the j^th^ FSA. The estimation of variability at different levels can be used to calculate ICC, which gives information about the proportion of the total variance in SARS-CoV-2 student infections that can be attributed to FSAs.

Model 2 includes predictors of SARS-CoV-2 student infections at the school level and area level. At the school level, low-income household, language mismatch, and low-parental education were included as predictors. At the area level, the four dimensions of the ON-Marg were included as predictors. School-level predictors are group-mean centered to remove between-area variations that would have otherwise been attributed to the school-level.^47^

Model 2 is as follows:

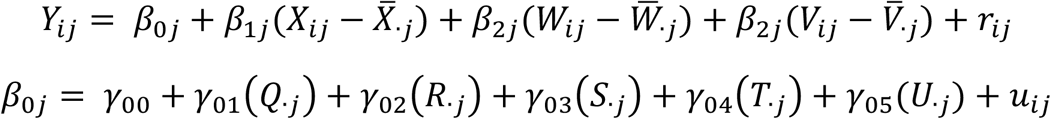

Where *Y*_*ij*_ refers to cumulative incidence of SARS-CoV-2 student infections for the i^th^ publicly funded elementary school in the j^th^ FSA. The predictors *Q*_.*j*_, *R*_.*j*_, *S*_.*j*_, *T*_.*j*_, *U*_.*j*_ refer to residential instability, material deprivation, dependency, ethnic concentration, and population size of the FSA, respectively.

## Appendix D. Additional Analysis

Model D1 includes the three school-level predictors (i.e., low-income households, language mismatch, low parental education) at the school level (as individual school values) and at the area level (as FSA means). Further, the population of the FSA is added as a predictor at the area level. We can estimate contextual effects by including FSA means of the three school-level predictors. For example, there is likely a contextual effect of language mismatch if the FSA means of language mismatch makes an independent contribution to predicting the cumulative incidence of SARS-CoV-2 student infections above and beyond what the individual school’s language mismatch value can predict.

Model D1 includes school characteristics (household income, language, and parental education) as predictors. At Level 1, the three variables were group-mean centered and entered as predictors. At Level 2, the aggregates (i.e., mean) of the three school characteristics were entered as predictors.^48^ Model 2 is as follows:

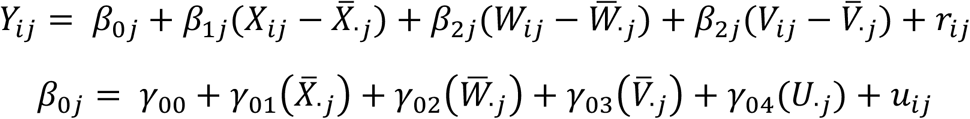

Where *Y*_*ij*_ refers to the cumulative incidence of SARS-CoV-2 student infections for the i^th^ publicly funded elementary school in the j^th^ FSA. The predictors *X*_*ij*_, *W*_*ij*_, and *V*_*ij*_ refer to low-income households, language mismatch, and low parental education, respectively. *U*_.*j*_ refers to the population of the FSA.

### Results

Model D examines the degree to which school-level variables and the aggregation of school-level variables at the FSA-level may predict the cumulative incidence of SARS-CoV-2 student infections (Table D1). Results indicate that higher proportions of low-income households at the school level and FSAs with schools with higher proportions of language mismatch at the FSA level significantly predict cumulative incidence.

There was a significant association between proportion of students from low-income households within schools and the cumulative incidence at the school level, but not at the area level.

However, the relationship at the school level was relatively weak, accounting for only 0.81% of the variability. In contrast, language mismatch was significantly positively associated with cumulative incidence at the area level but not at the school-level, suggesting that areas with high language mismatch were also areas with relatively higher cumulative incidence.

**Table D1.**
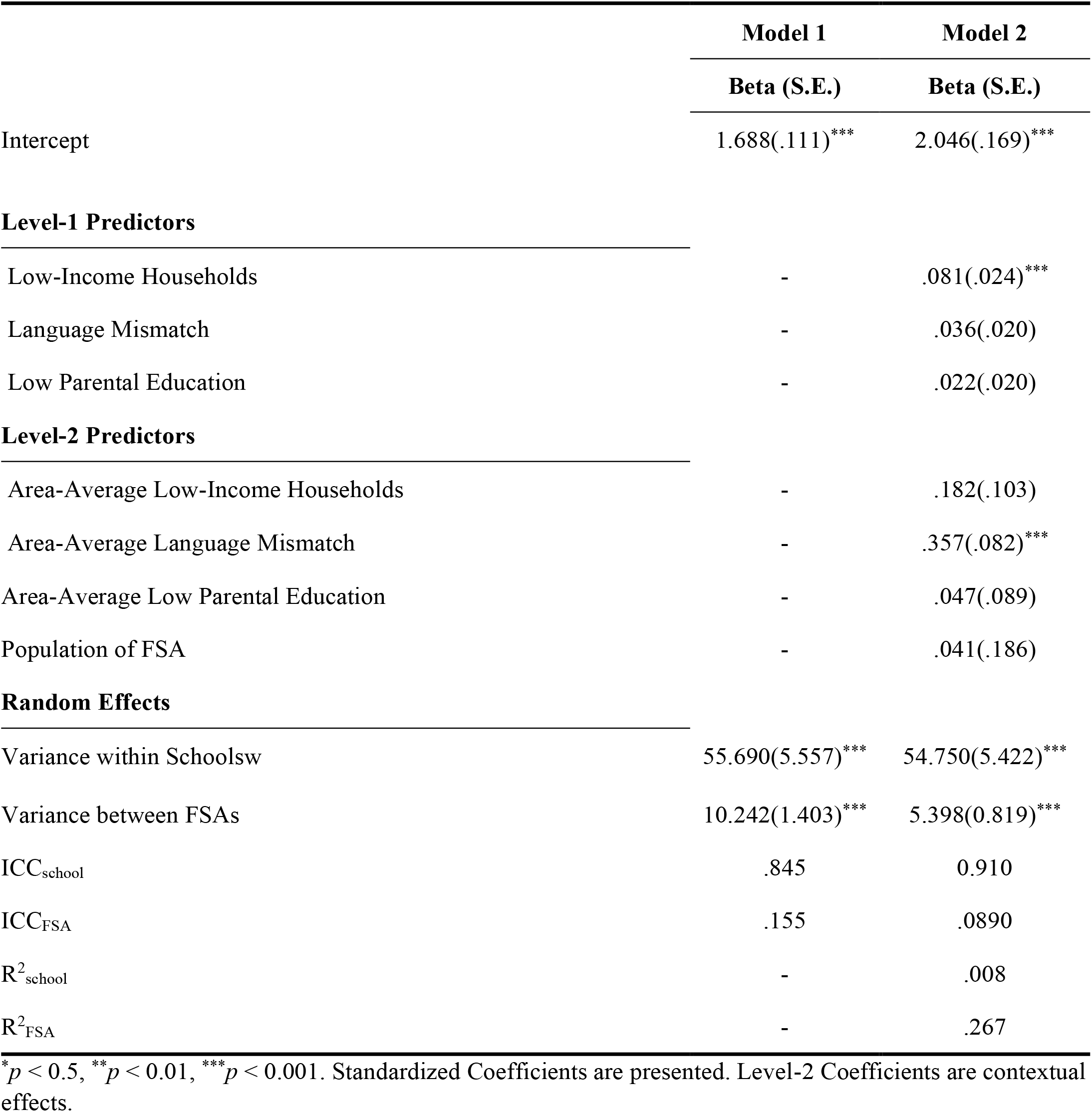
Multilevel Models

|  | **Model 1** | **Model 2** |
| --- | --- | --- |
|  | **Beta (S.E.)** | **Beta (S.E.)** |
| Intercept | 1.688(.111)<sup>***</sup> | 2.046(.169)<sup>***</sup> |
| **Level-1 Predictors** |  |  |
| Low-Income Households | - | .081(.024)<sup>***</sup> |
| Language Mismatch | - | .036(.020) |
| Low Parental Education | - | .022(.020) |
| **Level-2 Predictors** |  |  |
| Area-Average Low-Income Households | - | .182(.103) |
| Area-Average Language Mismatch | - | .357(.082)<sup>***</sup> |
| Area-Average Low Parental Education | - | .047(.089) |
| Population of FSA | - | .041(.186) |
| **Random Effects** |  |  |
| Variance within Schools<sub>w</sub> | 55.690(5.557)<sup>***</sup> | 54.750(5.422)<sup>***</sup> |
| Variance between FSAs | 10.242(1.403)<sup>***</sup> | 5.398(0.819)<sup>***</sup> |
| ICC<sub>school</sub> | .845 | 0.910 |
| ICC<sub>FSA</sub> | .155 | .0890 |
| R<sup>2</sup><sub>school</sub> | - | .008 |
| R<sup>2</sup><sub>FSA</sub> | - | .267 |

\* $p < 0.5$ , \*\* $p < 0.01$ , \*\*\* $p < 0.001$ . Standardized Coefficients are presented. Level-2 Coefficients are contextual effects.

## Appendix E. Sensitivity Analysis

One other possible method to operationalize the outcome variable is to use the cumulative number of cases per school rather than cumulative incidence. In addition to being a standard metric used in other studies,^49,50^ we believe cumulative incidence is the superior metric in the current context as it adjusts for the relative population of the school. To ascertain whether the use of cumulative cases rather than cumulative incidence might substantially alter the research findings of the current study, the main analyses were repeated with cumulative cases as the outcome variable.

Results are reported in Table E1. Results are very similar to the results reported in the main study and suggests that the research findings of the current study are robust to differences in how the independent variable is operationalized.

**Table E1.** Multilevel Models with Cumulative Cases as Dependent Variable

|  | Model 1 | Model 2 |
| --- | --- | --- |
|  | Beta (S.E.) | Beta (S.E.) |
| Intercept | 2.005(.095)*** | 1.921(.153)*** |
| Level-1 Predictors |  |  |
| Low-Income Households | - | .062(.025)* |
| Language Mismatch | - | .061(.016)*** |
| Low Parental Education | - | .024(.017) |
| Level-2 Predictors |  |  |
| Population of FSA | - | -.080(.064) |
| Residential Instability | - | .180(.054)** |
| Material Deprivation | - | .170(.075)** |
| Dependency | - | -.251(.052)*** |
| Ethnic Concentration | - | .507(.044)*** |
| Random Effects |  |  |
| Variance within Schools | 6.171(.659)*** | 6.391(.691)*** |
| Variance between FSAs | 2.945(1.397)*** | 1.249(.681)*** |
| ICC<sub>school</sub> | .678 | .837 |
| ICC<sub>FSA</sub> | .323 | .163 |
| R<sup>2</sup><sub>school</sub> | - | .011 |
| R<sup>2</sup><sub>FSA</sub> | - | .518 |

\*p < 0.5, \*\*p < 0.01, \*\*\*p < 0.001. Standardized Coefficients are presented.

## Footnotes

1 Choi, K. H., Denice, P., Haan, M., & Zajacova, A. (2021). Studying the social determinants of COVID-19 in a data vacuum. *Canadian Review of Sociology*, 58(2), 146–164. https://doi.org/10.1111/cars.12336

2 Choi, K. H., & Denice, P. (2020). Neighborhood SES and the COVID-19 pandemic. *SocArXiv*. https://doi.org/10.31235/osf.io/3xg5q

3 Fox, A. M., Lee, J. S., Sorensen, L. C., & Martin, E. G. (2021). Sociodemographic characteristics and inequities associated with access to in-person and remote elementary schooling during the COVID-19 pandemic in New York State. *JAMA Network Open*, 4(7), e2117267–e2117267. https://doi.org/doi:10.1001/jamanetworkopen.2021.17267

4 Donnelly, R., & Patrinos, H. A. (2021). Learning loss during Covid-19: An early systematic review. *Prospects*. https://doi.org/10.1007/s11125-021-09582-6

5 Gallagher-Mackay, K., Srivastava, P., Underwood, K., Dhuey, E., McCready, L., Born, K.B., Maltsev, A., Perkhun, A., Steiner, R., Barrett, K., & Sander, B. (2021). COVID-19 and education disruption in Ontario: Emerging evidence on impacts, (Ver. 1.1). Toronto: Ontario COVID-19 Science Advisory Table. https://doi.org/10.47326/ocsat.2021.02.34.1.0

6 Hammerstein, S., König, C., Dreisörner, T., & Frey, A. (2021). Effects of COVID-19-related school closures on student achievement - A systematic review. *Frontiers in Psychology*, 12. https://doi.org/10.3389/fpsyg.2021.746289

7 Kelly, A. B., O’Flaherty, M., Connor, J. P., Homel, R., Toumbourou, J. W., Patton, G. C., & Williams, J. (2011). The influence of parents, siblings and peers on pre- and early-teen smoking: A multilevel model. *Drug and Alcohol Review*, 30(4), 381–387. https://doi.org/10.1111/j.1465-3362.2010.00231.x

8 Merlo, J., Chaix, B., Ohlsson, H., Beckman, A., Johnell, K., Hjerpe, P., Råstam, L., & Larsen, K. (2006). A brief conceptual tutorial of multilevel analysis in social epidemiology: Using measures of clustering in multilevel logistic regression to investigate contextual phenomena. *Journal of Epidemiology and Community Health*, 60(4), 290. https://doi.org/10.1136/jech.2004.029454

9 Muijs, D. (2017). Can schools reduce bullying? The relationship between school characteristics and the prevalence of bullying behaviours. *British Journal of Educational Psychology*, 87(2), 255–272. https://doi.org/10.1111/bjep.12148

10 Government of Ontario. (2020a). 2018-2019 Academic year. Final as of September 4, 2020. Ontario public schools enrolment—Ontario Data Catalogue. Toronto: Queen’s Printer for Ontario. https://data.ontario.ca/dataset/ontario-public-schools-enrolment

11 Government of Ontario. (2020b). 2018-2019 Academic year. Final as of November 6, 2020. Private school enrolment by gender—Ontario Data Catalogue. Toronto: Government of Ontario. https://data.ontario.ca/en/dataset/private-school-enrolment-by-gender

12 Gallagher-Mackay et al., 2021.

13 Srivastava, P., & Taylor, P. J. (2021). COVID-19 school dashboard (1.1 May 2021) [Web application]. http://covid19schooldashboard.com/

14 Government of Ontario. (2021a). Ontario’s digital and data directive, 2021. [Website]. Toronto: Government of Ontario. https://www.ontario.ca/page/ontarios-digital-and-data-directive-2021

15 Government of Ontario. (2021d). Schools with recent COVID-19 cases. April 27, 2021 update. Toronto: Queen’s Printer for Ontario. https://data.ontario.ca/dataset/summary-of-cases-in-schools/resource/8b6d22e2-7065-4b0f-966f-02640be366f2 In earlier iterations on the Ontario Data Directive this dataset was named, *Schools with Active COVID-19 Cases* (April 27, 2021 update).

16 Government of Ontario. (2021b). School information and student demographics. June 29, 2021 update. Ontario Data Catalogue. Toronto: Queen’s Printer for Ontario. At the time of writing, this dataset was no longer available from the data archive. Researchers should contact the Ontario Data Catalogue if interested.

17 Matheson, F. I., & Van Ingen, T. (2018). 2016 Ontario Marginalization Index: User guide. Toronto: St. Michael’s Hospital/Public Health Ontario. https://www.publichealthontario.ca/-/media/documents/O/2017/on-marg-userguide.pdf

18 Statistics Canada. (2017b). Population and dwelling count highlight tables, 2016 census [Database]. Ottawa: Statistics Canada, Government of Canada. https://www12.statcan.gc.ca/census-recensement/2016/dp-pd/hlt-fst/pd-pl/Table.cfm?Lang=Eng&T=1201&S=22&O=A

19 Auger, K. A., Shah, S. S., Richardson, T., Hartley, D., Hall, M., Warniment, A., Timmons, K., Bosse, D., Ferris, S. A., & Brady, P. W. (2020). Association between statewide school closure and COVID-19 incidence and mortality in the US. *JAMA*, 324(9), 859–870. https://doi.org/doi:10.1001/jama.2020.14348

20 Rafael, R. de M. R., Neto, M., Depret, D. G., Gil, A. C., Fonseca, M. H. S., & Souza-Santos, R. (2020). Effect of income on the cumulative incidence of COVID-19: An ecological study. *Revista Latino-Americana de Enfermagem*, 28, e3344–e3344. https://doi.org/10.1590/1518-8345.4475.3344

21 Donnelly & Patrinos, 2021.

22 Hammerstein et al., 2021.

23 The School Information and Student Demographics dataset also included variables indicating whether students were new immigrants from non-English or non-French countries. However, this variable was not included as it was highly correlated with language mismatch, leading to multicollinearity concerns.

24 Schools in Ontario closed for spring break from 12-16 April 2021. Schools started emergency remote virtual instruction from 19 April 2021. They did not reopen for in-person instruction for the reminder of the school year ending June 2021.

25 Auger et al., 2020.

26 Rafael et al., 2020.

27 Statistics Canada. (2017a). Dictionary, census of population, 2016—Low-income measure, after tax (LIM-AT) [Website]. Ottawa: Statistics Canada, Government of Canada. https://www12.statcan.gc.ca/census-recensement/2016/ref/dict/fam021-eng.cfm

28 Government of Ontario. (2021c). School information finder [Website]. Toronto: Government of Ontario. https://www.app.edu.gov.on.ca/eng/sift/glossary.asp

29 FSAs with schools comprising junior kindergarten to Grade 6 (JK-6) and junior kindergarten to Grade 8 (JK-8) were included.

30 Population figures from the 2022 census were available. However, we used 2016 census data for FSA population size to increase compatibility with available ON-Marg data, which were derived from the 2016 census.

31 Marsh, H. W., Lüdtke, O., Nagengast, B., Trautwein, U., Morin, A. J. S., Abduljabbar, A. S., & Köller, O. (2012). Classroom climate and contextual effects: Conceptual and methodological issues in the evaluation of group-level effects. *Educational Psychologist*, 47(2), 106–124. https://doi.org/10.1080/00461520.2012.670488

32 All data and code for analysis in this paper is available via open access from: https://dataverse.scholarsportal.info/dataverse/COVID19SchoolInfections

33 Muthén, L. K., & Muthén, B. O. (2019). Mplus (version 8.3) [Statistical software]. Los Angeles.

34 Finch, J. F., West, S. G., & MacKinnon, D. P. (1997). Effects of sample size and nonnormality on the estimation of mediated effects in latent variable models. Structural Equation Modeling: A Multidisciplinary Journal, 4(2), 87–107. https://doi.org/10.1080/10705519709540063

35 Given this moderate departure from normality, the maximum likelihood estimator with robust standard errors is appropriate for model estimations.

36 Matheson & Van Ingen, 2018.

37 Public Health Ontario. (2021). COVID-19 in Ontario: Elementary and secondary school outbreaks and related cases, August 30, 2020 to April 24, 2021. Toronto: Queen’s Printer for Ontario. https://www.publichealthontario.ca/-/media/documents/ncov/epi/2020/12/covid-19-school-outbreaks-cases-epi-summary.pdf

38 Mishra, S., Ma, H., Moloney, G., Yiu, K. C. Y., Darvin, D., Landsman, D., Kwong, J. C., Calzavara, A., Straus, S., Chan, A. K., Gournis, E., Rilkoff, H., Xia, Y., Katz, A., Williamson, T., Malikov, K., Kustra, R., Maheu-Giroux, M., Sander, B., & Baral, S. D. (2022). Increasing concentration of COVID-19 by socioeconomic determinants and geography in Toronto, Canada: An observational study. *Annals of Epidemiology*, 65, 84–92. https://doi.org/10.1016/j.annepidem.2021.07.007

39 Chagla, Z., Ma, H., Sander, B., Baral, S. D., Moloney, G., & Mishra, S. (2021). Assessment of the burden of SARS-CoV-2 variants of concern among essential workers in the Greater Toronto Area, Canada. *JAMA Network Open*, 4(10), e2130284–e2130284. https://doi.org/10.1001/jamanetworkopen.2021.30284

40 Mishra et al., 2022.

41 Sundaram, M. E., Calzavara, A., Mishra, S., Kustra, R., Chan, A. K., Hamilton, M. A., Djebli, M., Rosella, L. C., Watson, T., Chen, H., Chen, B., Baral, S. D., & Kwong, J. C. (2021). Individual and social determinants of SARS-CoV-2 testing and positivity in Ontario, Canada: A population-wide study. *CMAJ*, 193(20), E723–E734. https://doi.org/10.1503/cmaj.202608

42 Husby, A., Corn, G., & Krause, T. G. (2021). SARS-CoV-2 infection in households with and without young children: Nationwide cohort study. *MedRxiv*. https://doi.org/10.1101/2021.02.28.21250921

43 Fox et al., 2021.

44 Donnelly & Patrinos, 2021.

45 Hammerstein et al., 2021.

46 Viner, R. M., Russell, S. J., Croker, H., Packer, J., Ward, J., Stansfield, C., Mytton, O., Bonell, C., & Booy, R. (2020). School closure and management practices during coronavirus outbreaks including COVID-19: A *Lancet Child & Adolescent Health*, 4(5), 397–404. https://doi.org/10.1016/S2352-4642(20)30095-X

47 Van den Noortgate, W., Opdenakker, M.-C., & Onghena, P. (2005). The effects of ignoring a level in multilevel analysis. *School Effectiveness and School Improvement*, 16(3), 281–303. https://doi.org/10.1080/09243450500114850

48 Enders, C. K., & Tofighi, D. (2007). Centering predictor variables in cross-sectional multilevel models: A new look at an old issue. *Psychological Methods*, 12(2), 121–138. https://doi.org/10.1037/1082-989X.12.2.121

49 Auger et al., 2020.

50 Rafael et al., 2020.

## Notes

### Competing Interest Statement

The authors have declared no competing interest.

### Funding Statement

This study did not receive any funding.

### Author Declarations

schoolsactivecovid.csv: https://data.ontario.ca/dataset/summary-of-cases-in-schools/resource/8b6d22e2-7065-4b0f-966f-02640be366f2 new_sif_data_table_2019_20prelim_en_june2021.xlsx: https://data.ontario.ca/dataset/school-information-and-student-demographics Population and Dwelling Count Highlight Tables, 2016 Census. : https://www12.statcan.gc.ca/census-recensement/2016/dp-pd/hlt-fst/pd-pl/Table.cfm?Lang=Eng&T=1201&S=22&O=A index-on-marg.xlsx: https://www.publichealthontario.ca/en/data-and-analysis/health-equity/ontario-marginalization-index

